# Screening for SARS-CoV-2 infection in asymptomatic individuals using the Panbio™ COVID-19 Antigen Rapid Test (Abbott) compared to RT-qPCR

**DOI:** 10.1101/2020.12.03.20243311

**Authors:** Beatrice M.F. Winkel, Emma Schram, Hendrik Gremmels, Sylvia B. Debast, Rob Schuurman, Annemarie M.J. Wensing, Marc J.M. Bonten, Edwin Goedhart, L. Marije Hofstra, Antigen Rapid Test Validation Group

## Abstract

**Background:** Antigen-based point of care tests for identification of SARS-CoV-2 may markedly enhance effectiveness of population-based controlling strategies. Previous studies have demonstrated acceptable sensitivity and high specificity compared to RT-qPCR in symptomatic individuals, but test performance for asymptomatic individuals is unknown.

**Methods:** Test performance of the Panbio™ COVID-19 Ag Rapid Test (Abbott) was compared to RT-qPCR in a longitudinal cohort study of asymptomatic football players and staff members of professional football clubs. Based on timing of symptoms and prior and subsequent test results, positive RT-qPCR tests were categorized as pre-symptomatic, early or late infection or persistent RNA shedding.

**Findings:** 2425 tests were performed in 824 individuals, of which 52 (6.3%) were SARS-CoV-2 positive based on RT-qPCR. There were 2406 paired sets from asymptomatic subjects for analysis. Sixteen Panbio™ tests were inconclusive, for which sensitivity analyses were performed (considering results as either positive or negative or being excluded). Sensitivity of Panbio™ ranged from 61.76% (95% CI 49.2-73.3) to 69.12% (95% CI: 56.7-79.8) and specificity from 99.53% (95% CI: 99.2-99.8) to 100% (95% CI: 99.8-100). Sensitivity of Panbio™ to detect subjects with pre-symptomatic/early infection (n= 42) ranged from 81.82% (95% CI: 67.3-91.8) to 90.91% (95% CI: 78.3-97.5) with specificity always above 99%.

**Interpretation:** In asymptomatic subjects the Panbio™ COVID-19 Ag Rapid Test had sensitivity of 81.82% to 90.91% and specificity above 99% for identifying pre-symptomatic and early SARS-CoV-2 infection.

**Funding:** This study was funded by the executing institutes. The Panbio™ COVID-19 Ag Rapid Tests were provided by the Ministry of Health, Welfare and Sport (VWS).

## Introduction

Rapid diagnosis of SARS-CoV-2 infection and subsequent contact tracing are essential in the containment of the current pandemic [1]. In most countries testing is targeted towards individuals with symptoms of a potential SARS-CoV-2 infection. However, infectiousness most likely occurs before symptom onset and some infections remain asymptomatic [2]. Screening of asymptomatic individuals may therefore also identify infectious individuals.

Currently, reverse transcriptase real time polymerase chain reaction (RT-qPCR) is the standard for detection of SARS-CoV-2 infection. In a validation study of the Abbott Panbio™ COVID-19 Ag rapid test in community-dwelling subjects with symptoms of respiratory tract infection, this assay had specificity of 100%, and sensitivity above 95% to detect SARS-CoV-2 infected subjects with low Ct-values in RT-qPCR (i.e. infections with a high viral load in nasopharyngeal samples) [3]. Considering the short turnaround time, user friendliness, low cost, and opportunity for decentralized testing, lateral flow assays (LFA) would also be a suitable test for screening of asymptomatic individuals.

We, therefore, validated the Panbio™ COVID-19 Ag rapid test in a cohort of asymptomatic football players and staff of football clubs from the Dutch national leagues who are longitudinally tested for SARS-CoV-2 infection with RT-qPCR at approximately weekly intervals.

## Materials and methods

### Populations and Study Period

The study population consisted of football players, staff and referees from 13 different professional football clubs and the national teams in the Netherlands. Starting August 2020, the Royal Netherlands Football Association (KNVB) required all individuals to be tested for SARS-CoV-2 infection by RT-qPCR independent of presence of symptoms, 2 days prior to each match. The study period of the LFA validation ran from 1^st^ of October, 2020 to 9^th^ of November 2020. All individuals were sampled for routine RT-qPCR testing, using a throat/nasopharyngeal swab, and underwent an additional nasopharyngeal swab for LFA testing. At two test visits for two different teams (n= 104 tests) only LFA tests were performed, followed by sampling for RT-qPCR on the following day. Data regarding symptoms of potential SARS-CoV-2 infection before and during the study period and (prior) test results were collected by the physicians of each of the football clubs.

### Diagnostic tests

#### RT-qPCR

The samples were tested with RT-qPCR by either Eurofins (Brugge, Belgium), Synlab Laboratories (Luik, Belgium), or U-diagnostics (Utrecht, the Netherlands). RT-qPCR results were obtained from the physicians of the participating football clubs and positive or discrepant results were verified. Residual material of these samples was not available for further analyses or repeated testing on a single platform.

#### LFA

The Panbio™ COVID-19 Ag rapid test device by Abbott (Lake Country, IL, U.S.A) is a membrane-based immunochromatography assay which detects the nucleocapsid protein of SARS-CoV-2 in nasopharyngeal samples. Collected swabs were transferred into dedicated sample collection tubes containing a lysis buffer provided with the test kit. Samples were processed on site, directly after collection, by trained personnel wearing personal protective equipment and in accordance with the manufacturer’s protocol. Test results were recorded after 15 minutes of assay initiation and documented by photograph.

### Analysis

Specificity and sensitivity with 95% confidence intervals of the LFA were calculated using the RT-qPCR results as reference test. The RT-qPCR results were categorized into different phases of infection. Presymptomatic infection was defined as a first positive test before onset of symptoms. Early infection includes the first positive test for individuals with asymptomatic infection (and therefore not identified as presymptomatic) and all positive test results within 7 days of the first positive test. Late infection was defined as a positive test at least 7 days after the first positive RT-qPCR result or at least 7 days after onset of symptoms with relief of symptoms for more than 24 hours. Persistent RNA shedding was defined as a positive test more than four weeks after the first positive RT-qPCR result and not suspected for re-infection based on absence of symptoms and high Ct values (>32). The LFA results were analysed for these different phases. Specificity and sensitivity were determined based on presymptomatic/ early infection versus late infection/ no infection. Since RT-qPCR was performed in different laboratories and using different assays, Ct values cannot directly be compared. Therefore no sensitivity analyses based on Ct values were performed.

### Ethical Approval

The medical research ethics committee (MREC) of Utrecht decided the study is not subject to the Medical Research Involving Human Subjects Act (WMO) and did not require full review by an accredited MREC. All participants have provided written informed consent.

## Results

### Population Characteristics

Our cohort included 824 individuals (94% male) with a median age of 27 years (range 16 to 80 years, IQR: 21-40). Overall, 2425 samples were tested by RT-qPCR and LFA. Of these, 2321 tests were obtained at the same time, and 104 tests were taken with a one day interval between LFA and PCR. The median number of LFA tests per subject was 3 (range 1-6, IQR: 1-4).

Based on RT-qPCR, 52 of 824 subjects tested positive for SARS-CoV-2 at least once during the study period (prevalence of SARS-CoV-2 infection: 6.3%). Of these 52, 29 developed (mostly mild) symptoms of SARS-CoV-2 infection; coryza (n=12), sore throat (n=12), fever (n=12), headache (n=6), muscle ache (n=5), cough (n=5), altered smell or taste (n=5). An asymptomatic infection occurred in 23 individuals (44%; prevalence of asymptomatic SARS-CoV-2 infection: 2.8%).

Of the 2425 tests, 18 were performed while the individual reported symptoms at time of testing; either at time of the first positive test (n=13) or during a follow-up after a prior positive test (n=5). These 18 tests were not included for analysis. One LFA test was invalid due to absence of a control band and was excluded for analysis. The comparison between RT-qPCR and LFA, therefore, includes 2406 paired tests.

### LFA results

Of 2406 tests, 68 (2.8%) were positive by RT-qPCR and 42 (1.7%) by LFA (Table 1). Sixteen LFA tests (0.67%) were considered inconclusive because of weak unclear bands, of which 5 were positive with RT-qPCR. We, therefore, determined sensitivity of LFA by either excluding them from analysis, or by considering them as positive or negative to determine best case and worst case scenarios. Sensitivity of LFA ranged from 69.12% (95% CI: 56.7-79.8) with inconclusive test results considered positive to 61.76% (95% CI: 49.2-73.3) with inconclusive test results considered negative. Specificity of LFA was 99.53% (95% CI: 99.2-99.8) with inconclusive test results considered positive and 100% (95% CI: 99.8-100) with inconclusive test results considered negative or excluded.

**Table 1:**
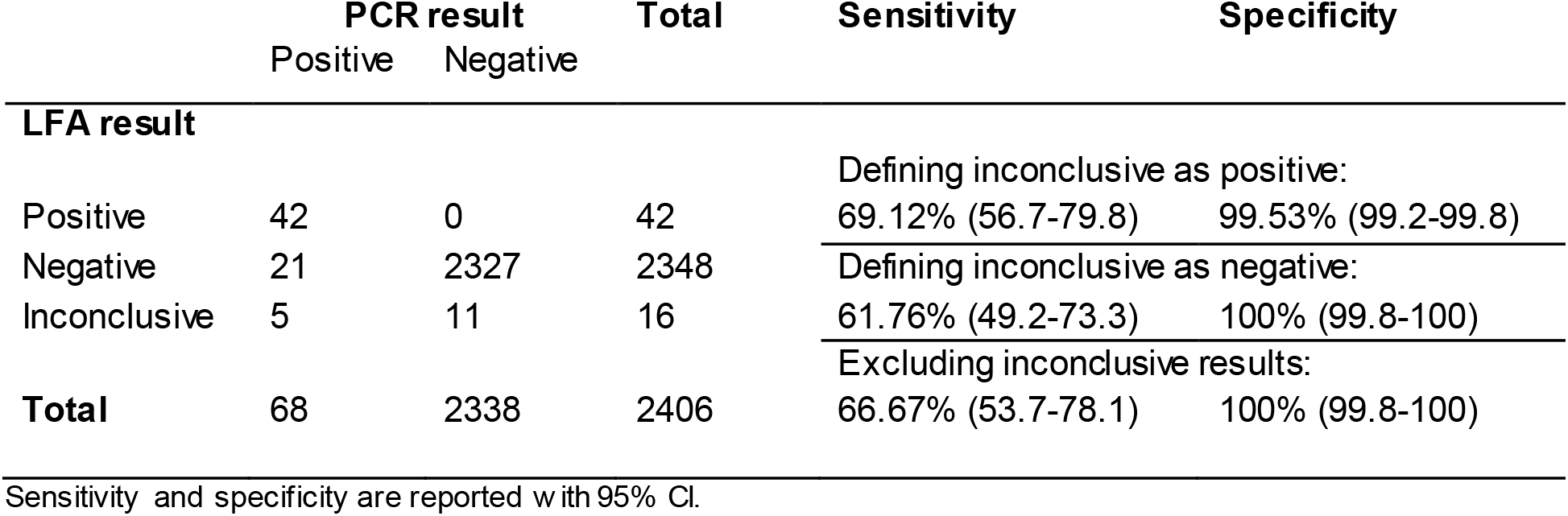
Test characteristics of the LFA compared to the RT-qPCR.

The positive RT-qPCR results were classified as presymptomatic (n=12), early infection (n=32), late infection (n=21), and persistent RNA shedding (n=3), and test characteristics were determined for presymptomatic/early infection, late infection/ persistent RNA shedding and no infection (Table 2). Sensitivity of LFA to detect subjects with presymptomatic/early infection ranged from 81.82% (95% CI: 67.3-91.8) with inconclusive test results considered negative, to 90.91% (95% CI: 78.3-97.5) with inconclusive test results considered positive, with specificity always above 99%. Sensitivity of LFA to detect subjects with late infection/ persistent RNA shedding ranged from 25.0% (95% CI: 9.8-46.7) to 29.17% (95%CI: 12.6-51.1), with specificity always above 99%.

**Table 2:**
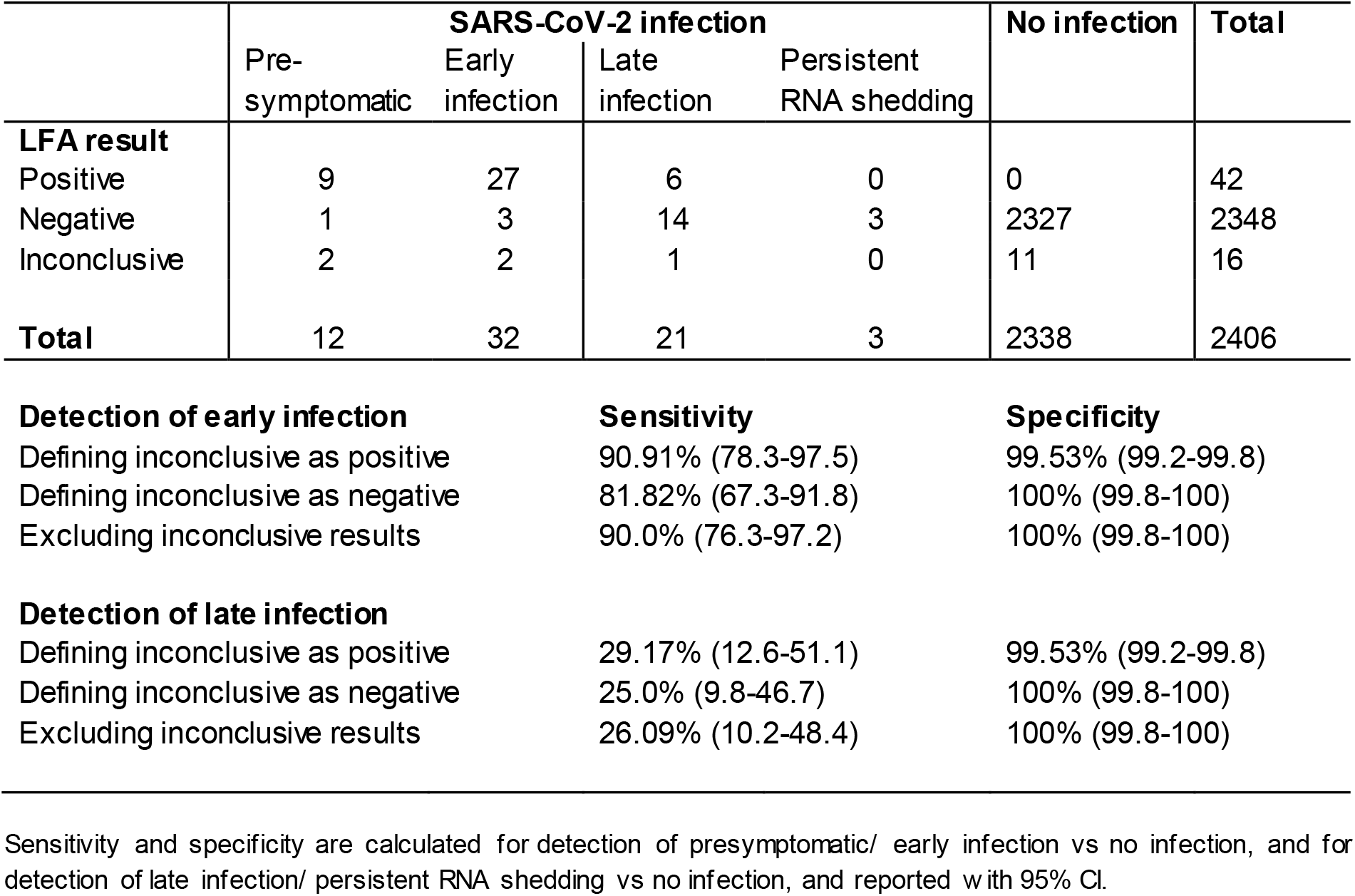
Test characteristics of the LFA for detection of early and late SARS-CoV-2 infection.

### Discrepancy analysis

False positive LFA results were not observed. Eleven LFA tests, obtained from 11 individuals at distinct test visits, were considered inconclusive and were tested negative with RT-qPCR. Follow-up tests confirmed the negative RT-qPCR test result.

False negative LFA results were observed for four individuals with presymptomatic or early infection. Two of them had Ct values of 27 and 29, and SARS-CoV-2 infection was confirmed in a subsequent test. In one of these subjects there was a sampling delay of one day between LFA and subsequent RT-qPCR. The other two false-negative tests occurred in asymptomatic individuals with a single positive RT-qPCR test with Ct values of 32 and negative RT-qPCR tests in the preceding and following week.

Most false negative LFA tests (17/21) occurred in subjects with late infection or persistent RNA shedding. All samples had Ct values above 27 (median Ct value 32 (IQR: 30-36)) and were obtained a median of 15 days (IQR: 10-18) after the first positive test. Compared to the false negative LFA tests during late infection, samples with a positive LFA result (6/21) had lower Ct values (median 27 (IQR: 22-28)) and were obtained a median of 10 days (IQR: 7 -11) after the first positive test.

## Discussion

The Panbio™ COVID-19 Ag rapid test reliably identified SARS-CoV-2 infected subjects in a cohort of asymptomatic individuals, with a specificity above 99% and an overall sensitivity of 69.12%. False negative LFA results were mostly observed in a late phase of infection (on average 2 weeks after the first positive test) with low viral loads in nasopharyngeal swabs. The estimated sensitivity of LFA in presymptomatic and early infections was between 81.82% and 90.91%.

Although our cohort is not a representative sample from the general population, as it constitutes predominantly males who are often young professional athletes, our results are in line with previous reports of this LFA. In our previous validation in an ambulant symptomatic population, overall sensitivity was 72.6% but increased to 95.2% when using a Ct-value of 32 as cut-off [3]. In a Spanish cohort of 1369 individuals an overall sensitivity of 71.4% was reported, which increased for symptomatic patients who presented within 5 days since symptom onset (83.1%), and for those with high viral load (87.7%) [4]. Another Spanish study retrospectively analysing frozen swabs reported a sensitivity of 79.5% among asymptomatic individuals and also observed an association with Ct-value (Ct-value <25 100%, Ct value <30 98.6%) [5].

For our analysis, three different laboratories performed the RT-qPCR analyses, limiting the possibility for overall analyses using Ct-values as a cut-off. The unique longitudinal prospective screening of this cohort enables us to distinguish different phases of infection. Th e Panbio™ COVID-19 Ag rapid test identified 11 of 12 presymptomatic infections and 29 of 32 early infections. In a longitudinal cohort of basketball players and staff, asymptomatic individuals rapidly progressed from a negative test to a peak Ct-value, as demonstrated by repeated testing [6], which could explain the good performance of this LFA during early infection. Sensitivity of the Panbio™ COVID-19 Ag rapid test decreased substantially during late infection (i.e. more than one week after the first positive test).

The Panbio™ COVID-19 Ag rapid test did not detect 14 of 21 late infections. Although samples were tested on different platforms limiting comparisons, all these samples had high Ct values. The risk of transmission is considered to be low or absent during late infection and will only further decrease in following days. Therefore, isolation of these individuals at that time most likely has limited effects from a public health perspective. This is supported by an analysis of LFA, RT-qPCR and virus culture, in which the Panbio™ COVID-19 Ag rapid test detected 97.3% of samples with positive cultures, reflecting infectious individuals [7].

Furthermore, modelling studies have demonstrated that surveillance effectiveness depends largely on frequency of testing and speed of reporting, rather than test sensitivity [8, 9]. The current study also demonstrates the feasibility of performing the Panbio™ COVID-19 Ag rapid test as point of care test, outside of a laboratory setting, by trained personnel with appropriate infection prevention control measures and use of personal protective equipment. Results were immediately reported to the physician of the football club, allowing direct intervention in case of positive LFA test results.

We observed inconclusive results in 0.66% of LFA tests. Yet, in five of these 16 inconclusive results SARS-CoV-2 appeared detectable with RT-qPCR. We, therefore, recommend additional sampling for RT-qPCR in case of inconclusive LFA results.

In conclusion, the Panbio™ COVID-19 Ag rapid test reliably identifies (early) SARS-CoV-2 infections in asymptomatic individuals, and can be used in targeted screening strategies for early detection of SARS-CoV-2 infection in asymptomatic individuals.

## Data Availability

De-identified participant data can be made available upon request per email to the corresponding author following publication.

## Author contributions

LMH and BW designed the study. MB, AW, RS, EG provided counsel on study design. Sample collection was performed by BW, ES, HG, SD and LMH. Data analysis was performed by BW, ES and LMH, and discussed with MB, AW, RS, HG, and EG. BW and LMH drafted the manuscript, and all authors contributed to critical revision of the manuscript.

## Acknowledgements

We thank all study participants from ADO Den Haag, AZ, Excelsior, FC Utrecht, Go Ahead Eagles, Heracles Almelo, KNVB, PEC Zwolle, PSV, RKC Waalwijk, SC Cambuur, Sparta, Telstar, for their commitment to take lessons from sports to enhance public health.

We also thank Ellen Broekhuizen, Lieke Broekmeulen, Alyssa de Bruijn, Wout Hamelink, Samantha Herrewegh, Priscilla Imthorn, Bente de Jager, Jens de Jager, Nynke de Jager, Lisanne Jannink, Omar Rogouti, Daniel Stieber, Anniek Tanja and Eva Theeuwes, and personnel of Eurofins, Synlab, and U-diagnostics who have assisted with collection and processing of samples.

